# Clonal expansion of cytotoxic CD8^+^ T cells in lecanemab-associated ARIA

**DOI:** 10.1101/2025.09.17.25335728

**Authors:** Lance A. Johnson, Kai Saito, Akhil V. Pallerla, Jessica L. Funnell, Ashley R. Ezzo, Chelsea L. Song, Douglas A. Harrison, Noah J. Norton, Lauren C. Moore, Linda J. Van Eldik, David W. Fardo, Greg E. Cooper, Josh M. Morganti

## Abstract

Amyloid-related imaging abnormalities (ARIA) remain the principal safety concern limiting adoption of anti-amyloid therapies such as lecanemab, yet their underlying biology is poorly defined. To address this, we performed deep multi-omic profiling of peripheral blood mononuclear cells from three Alzheimer’s disease (AD) patients who developed ARIA and three matched controls. Single-cell RNA sequencing, CITE-seq, V(D)J clonotyping, and metabolomic/lipidomic profiling revealed a coordinated reprogramming of the CD8^+^ compartment in ARIA+ patients. CD8^+^ TEM and TEMRA subsets were numerically expanded, transcriptionally enriched for cytotoxic and migratory programs, and exhibited increased clonal expansion. Transcription factor inference and metabolomics converged on a glycolytic bias, supporting short-lived effector activity. Ligand–receptor modeling identified ARIA-associated signaling from CD14^+^ and CD16^+^ monocytes that augmented antigen presentation, adhesion, and chemokine axes directed toward effector CD8s. Finaly, integration with an external cerebrovascular atlas confirmed that ARIA-associated TEM/TEMRAs are transcriptionally “addressed” for vascular engagement. Together, these findings establish a peripheral immune–vascular axis linking immunometabolic reprogramming, clonal cytotoxic CD8^+^ expansion, and altered monocyte signaling to ARIA, with implications for biomarker development and risk mitigation during anti-amyloid therapy.

**Significance:** Anti-amyloid therapies improve Alzheimer’s disease outcomes but are constrained by ARIA, a serious immune–vascular complication with unclear etiology. By integrating single-cell, clonotype, and metabolomic profiling, we show that ARIA is associated with glycolysis-driven expansion of cytotoxic CD8 TEM/TEMRA subsets that engage monocyte and endothelial signaling axes. These findings identify a peripheral immune program that may inform biomarker development and therapeutic strategies to mitigate ARIA risk.

## Main

Alzheimer’s disease (AD) is the leading cause of dementia worldwide, accounting for 60-80% of all documented dementia cases. In 2023, the first disease modifying therapeutic was fully approved by the FDA for use in treatment of AD. Lecanemab (Leqembi®), an anti-amyloid beta (Aβ) monoclonal antibody selective for soluble protofibrils, is remarkably efficient in clearing amyloid from the brain, and modestly slows the cognitive decline associated with AD ^1^. However, treatment is sometimes accompanied by amyloid-related imaging abnormalities (ARIA), including vasogenic edema/effusions (ARIA-E) and microhemorrhages/siderosis (ARIA-H) ^1,2^. With safety a key determining factor for both patients and providers and a high burden of MRI-based monitoring, access to and adoption of anti-amyloid therapies have been slowed by concerns over ARIA risk.

Susceptibility to ARIA is not uniform and its underlying mechanism(s) remain unknown. Current data indicate a strong Apolipoprotein E (*APOE*) ε4 gene-dose effect on ARIA incidence, motivating pre-treatment genotyping and intensified MRI monitoring in ε4 carriers ^3-5^. Case reports describe hemorrhages after thrombolysis^6^ and fatal β-amyloid–related arteritis^7^, both cases in *APOE* ε4/ε4 individuals. Collectively, these observations implicate an immune–vascular mechanism for ARIA and raise the question of whether circulating peripheral blood mononuclear cell (PBMC) signatures can forecast ARIA risk and clarify its biology. Because lecanemab is delivered systemically and ARIA risk peaks early, dynamic immune responses in the blood are both biologically relevant to cerebral events and practically measurable at the time when risk is greatest.

In this report, we describe a case-control study of three sex, age, and *APOE* genotype matched pairs of AD patients undergoing treatment with lecanemab at a regional medical center ^8^. Using next-generation sequencing techniques, including single-cell (sc) RNA sequencing (seq), CITE-seq, V(D)J, and targeted metabolomics and lipidomics, we identify a unique immunometabolic profile of subjects who develop ARIA, compared to those who do not. Specifically, PBMCs from ARIA+ patients feature expanded CD8^+^ T cell subsets and increased clonal expansion of CD8^+^ T-Effector Memory expressing CD45RA (TEMRA) cells. These ARIA+ PBMCs are transcriptionally distinct, enriched for pro-glycolytic genes and metabolites, and spatially map to brains of AD patients treated with active and passive Aβ immunization ^9^. Together, these clinical data highlight a novel T-cell mediated mechanism that may inform future studies and aid in the development of safer and more effective therapeutics for the highest risk AD populations.

## Results

### Expansion of cytotoxic CD8 subsets defines the ARIA immune signature

Six individuals receiving bi-weekly infusions (10 mg/kg) of lecanemab were recruited from Norton Neuroscience Institute Memory Clinic (Louisville, KY) ^8^. We selected three subjects with radiographically confirmed ARIA, including one case of ARIA-E+, one ARIA-H+, and one ARIA E+/H+ case (Fig. 1a). Controls were matched by age (± 2-5 years), sex, *APOE* genotype, and lecanemab infusion number (± 1); none of the individuals were taking additional immunotherapies (Supplemental Fig. 1). To establish the clinical context of our cohort, we first confirmed the presence of amyloid-related imaging abnormalities (ARIA) on serial MRI scans. Representative cases demonstrate ARIA-E, detected on T2-weighted fluid-attenuated inversion recovery (FLAIR/TIRM) imaging, and/or ARIA-H, detected on susceptibility-weighted imaging (SWI) (Fig. 1a).

**Figure 1.**
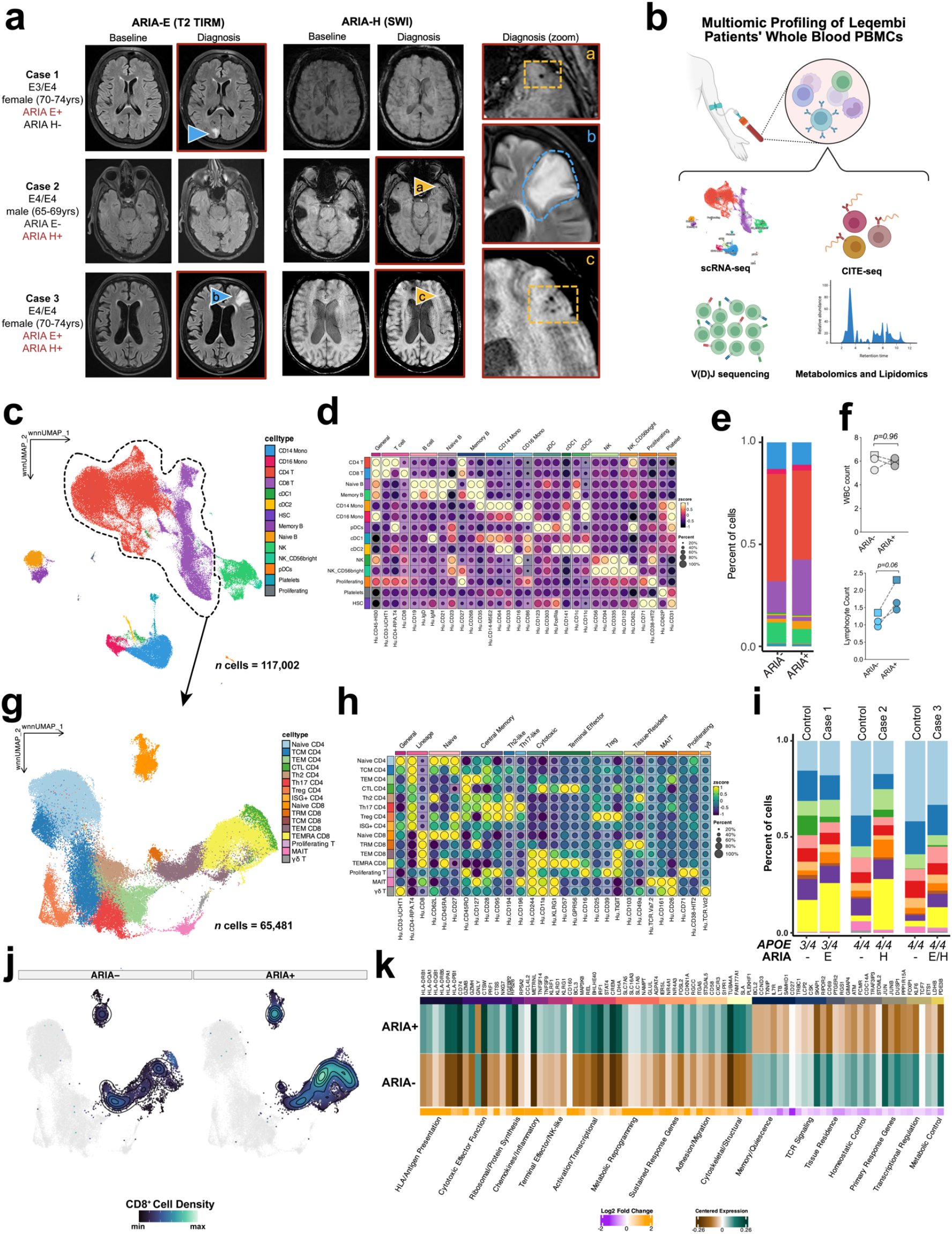
ARIA is associated with expansion of peripheral CD8^+^ TEMRA cells. **a)** Representative MRIs show edema/e5usions on T2-FLAIR/TIRM (blue triangles) and microhemorrhages/siderosis on SWI (orange triangles) from three selected ARIA cases (ARIA-E, ARIA-H, and ARIA-E/H). Cases were matched to three controls by age, sex, *APOE* genotype, and infusion number. **b)** Study design and multi-omic profiling workflow. PBMCs collected pre-infusion underwent 5′ scRNA-seq with CITE-seq and V(D)J, plus targeted metabolomics and lipidomics. **c–d)** Multimodal (RNA + Antibody-Derived Tags (ADT)) UMAP of >117,000 cells resolves major PBMC lineages (T, B, NK, monocytes, DCs). **e)** Proportions of major lymphocyte classes show increased CD8^+^ and decreased CD4^+^ frequencies in ARIA+ subjects. F) Routine clinical counts reveal similar CBC di5erentials across groups, with a trend toward higher absolute lymphocyte counts in ARIA+ cases. **g–h)** Re-clustering of 65,481 T cells identifies canonical CD4^+^ (naïve, central/e5ector memory, Tregs) and CD8^+^ (naïve, central/e5ector memory, TEMRA, intermediate cytotoxic) states using concordant RNA and surface markers. **i–j)** ARIA+ subjects exhibit contraction of naïve CD8^+^ cells and enrichment of e5ector memory/TEMRA subsets. Density plots (j) highlight TEMRA accumulation in ARIA+ only. **k)** Di5erential expression (Fisher meta-analysis) in TEMRA cells from ARIA+ versus ARIA-reveals increased cytotoxic/Fc-receptor signaling, chemokine/migratory genes, MHC-II, inhibitory/stress markers, and metabolic regulators consistent with a glycolytic, glutamine-fueled e5ector state.

To capture the peripheral immune repertoire, we performed multi-omic profiling of PBMCs from ARIA case-controls using single-cell RNA sequencing, CITE-seq, V(D)J clonotyping, and metabolomic/lipidomic profiling (Fig. 1b). Unsupervised clustering (integrated on RNA and surface protein expression) revealed the expected diversity of immune lineages across T lymphocytes (CD3^+^CD4^+^ and CD3^+^CD8^+^), B lymphocytes (CD19^+^), monocytes (CD14^+^ and CD16^+^), plasmacytoid (FcϵR1α^+^) and conventional (CD11c^+^) dendritic cells, natural killer cells (CD56^+^) and platelets (CD62p)(Fig. 1c-d). Examining proportional changes across these cell types revealed an overall increase in the amount of CD8^+^ T cells in the ARIA^+^ samples (Fig. 1e; purple) with a concomitant decrease in CD4^+^ T cells (Fig. 1e; red). Similarly, clinical blood work showed no difference in red or white blood cell counts, although an absolute lymphocyte count suggested an increase in ARIA+ subjects (Fig. 1f; Supplemental Fig. 1).

To examine proportional shifts beyond the generalized CD4 versus CD8 dichotomization, we subsetted and re-clustered CD4^+^ and CD8^+^ T cells to reveal 65,481 cells across 16 states in the integrated RNA + surface protein latent space (Fig. 1g). Cell-surface protein expression was used to corroborate RNA signatures and define the 16 states (Fig. 1h). Within the CD8^+^ compartment, comparative analysis showed a relative expansion of naïve cells (Naive; CD45RA^+^, CD62L^+^; CCR7^+^, SELL^+^), effector memory (TEM; CD45RO^+^, GZMA^+^, CCL5^+^), with the largest proportional increases observed in terminally differentiated effector memory (TEMRA; CD45RA^+^, KLRG1^+^, GNLY^+^, GZMB^+^)(Fig. 1i-j).

To evaluate whether these compositional changes were accompanied by transcriptional differences, we performed Fisher’s meta-analysis comparing TEMRAs from ARIA+ versus ARIA-samples (Fig. 1k). TEMRA cells from ARIA^+^ patients displayed a coordinated effector program: upregulated cytotoxic/Fc-receptor signaling (STX11, FCGR3A, LYN/FGR/PLCG2) and chemokine/migratory machinery (CCL3, CYRIA, SLC16A3) favoring vascular trafficking and sustained activity. The TEMRA cells from ARIA+ patients also expressed MHC-II genes (HLA-DRB1/-DQA1/-DQB1/-DRB5) and inhibitory/stress transcripts (LILRB1, CISH, CDKN1A, NR4A1), consistent with chronic antigenic stimulation alongside effector programming. These findings echo TEMRA biology in aging/chronic inflammation and multiple sclerosis (MS), where clonally expanded CD8^+^ TEMRA/Tissue Resident Memory (TRM) cells accumulate at perivascular/meningeal sites and can injure endothelium ^10-12^. Together, these results indicate that TEMRAs in ARIA+ patients adopt a unique transcriptional profile marked by effector function, migratory capacity, chronic activation, and a shifted metabolic signature.

An additional feature of ARIA+ TEMRAs was enrichment of metabolic regulators (down: LDHB; up: GLUL, LDHA, and the lactate and glutamine exporters SLC16A3 and SLC1A5) indicating a pro-glycolytic, glutamine-fueled metabolism that supports sustained effector activity (Fig. 1k) ^13^. To further explore whether the transcriptional differences identified in ARIA+ CD8^+^ subsets were linked to metabolic regulation, we first inferred transcription factor activity from our scRNA-seq dataset using decoupleR (Fig. 2a). This analysis highlighted shifts in several regulators of T cell metabolism, including consistent downregulation of VHL in ARIA+ TEM and TEMRA cells. VHL normally constrains hypoxia-inducible factor (HIF) signaling; its reduced activity in ARIA+ effector cells therefore suggests a potential stabilization of HIF-1α and a shift toward glycolytic metabolism, a state known to favor short-lived effector function over long-term persistence ^10,11^. Additional TF changes included modulation of PPARA, which regulates fatty acid oxidation, and altered activity of histone deacetylases (HDAC1/3), together indicating broad remodeling of metabolic and epigenetic programs.

**Figure 2.**
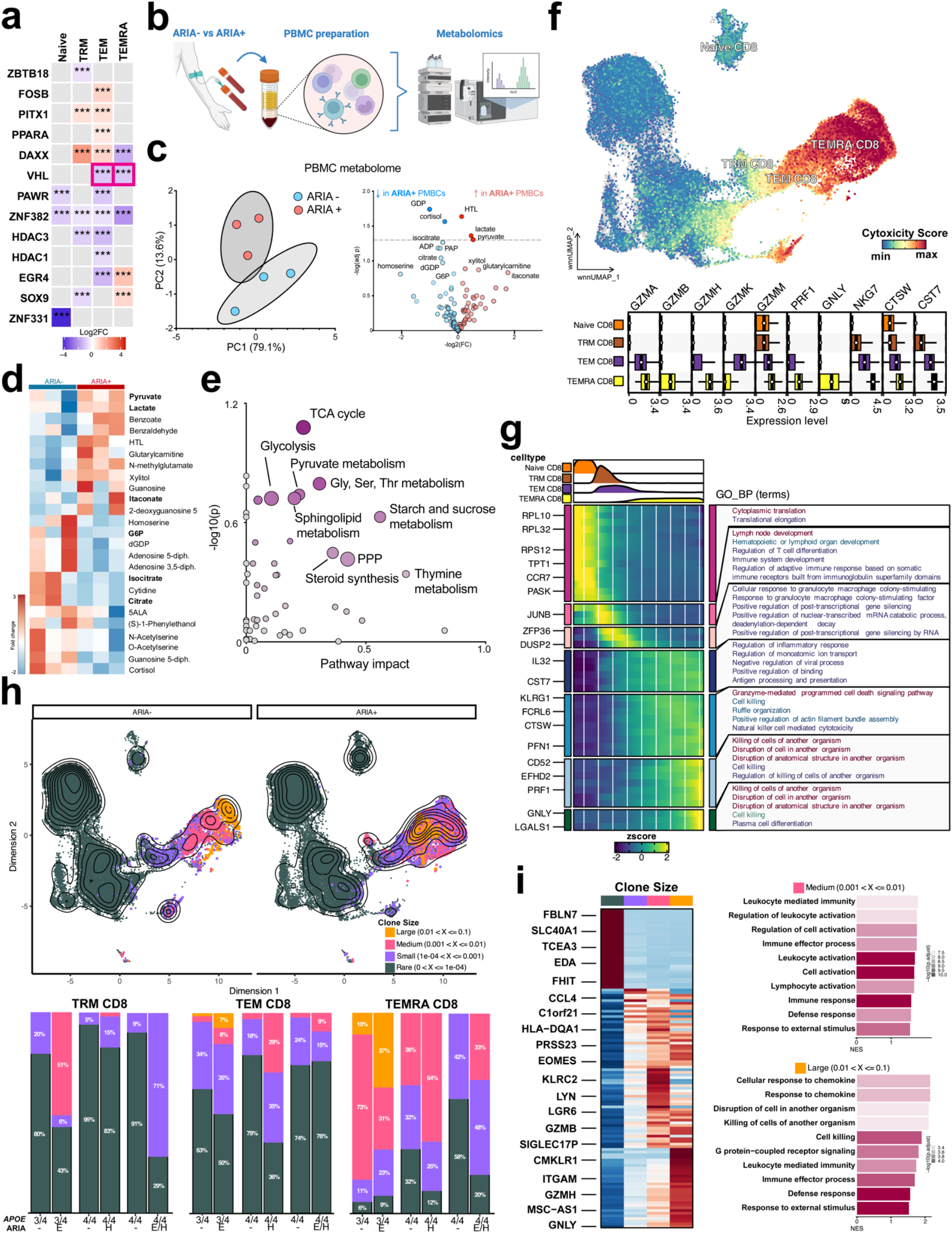
ARIA is associated with immunometabolic reprogramming, increased cytotoxicity, and clonal remodeling of CD8^+^ T cells. **a)**Transcription-factor activity inferred with decoupleR shows reduced VHL activity (red boxes) and altered PPARA/HDAC1/3 in ARIA+ TEM/TEMRA cells, consistent with HIF-1α stabilization and a shift toward glycolysis. **b)** Targeted PBMC metabolomics workflow. **c-d)** Principal-component analysis separates ARIA^+^ and ARIA^−^ samples. Di5erential metabolites within the volcano plot and heatmap highlight increased lactate, pyruvate, and itaconate with decreased citrate in ARIA+. **e)** Pathway impact analysis indicates ARIA-dependent shifts in glycolysis, pyruvate and sphingolipid metabolism, TCA, and pentose-phosphate pathways. **f)** Cytotoxicity module scores (top) and gene-level summaries (bottom) rise from naïve CD8^+^ through TRM-like and TEM to TEMRA states. **g)** Pseudotime from naïve CD8^+^ to TEMRA reveals early translational, intermediate signaling, and terminal granzyme-mediated cytotoxic programs. **h)** Clonal architecture shows a higher burden of medium/large clones in ARIA+ overall and per subject. **i)** Subset-wise clonality: naïve CD8^+^ unchanged; TRM-like exhibit selective small/medium expansions; TEM and TEMRA increase in both frequency and clonality in ARIA+. **j)** Larger clones are enriched for e5ector/cytotoxic genes (e.g., GZMB, GZMH, GNLY, CCL4, EOMES, LYN), NK-associated receptors (KLRC2, CMKLR1, SIGLEC17P), and antigen-presentation/adhesion (HLA-DQA1, ITGAM). **k)** Pathway analysis confirms progressive enrichment of cell-killing/cytotoxic e5ector programs with increasing clone size.

### Immunometabolic reprogramming fuels effector and clonal remodeling

To confirm these transcriptional inferences at the metabolite level, we performed targeted metabolomic profiling of PBMCs from ARIA+ and ARIA-patients (Fig. 2b). Principal component analysis revealed a clear separation of ARIA+ from ARIA-samples, demonstrating global differences in metabolite abundance (Fig. 2c, left). This separation was driven by several differentially abundant metabolites in ARIA+ samples, including increases in lactate, pyruvate and itaconate and decreases in citrate, characteristic of the ‘broken’ TCA cycle observed in pro-infiammatory myeloid cells (Fig. 2c-d)^12^. Pathway impact analysis further supported this observation, identifying enrichment of glycolysis, pyruvate metabolism, and sphingolipid metabolism, alongside perturbations in the TCA cycle and the pentose phosphate pathway (Fig. 2e). These ARIA-driven metabolic shifts are consistent with prior literature establishing a metabolic reprogramming inherent in the regulation of CD8^+^ effector states and heightened effector activity ^13-15^. Given that glycolytic reprogramming is closely coupled to effector function, we next examined whether ARIA-associated TEM and TEMRA cells also displayed evidence of augmented cytotoxic potential and clonal remodeling.

A cytotoxicity module score was lowest in CD4 populations and naïve CD8 cells, but progressively increased in TRM-like, TEM, and TEMRA subsets (Fig. 2f, top). Gene-level summaries demonstrated that cytotoxic mediators rose in a consistent stepwise manner across these states (Fig. 2f, bottom). To capture this trajectory more comprehensively, we applied pseudotime analysis seeded in naïve CD8^+^ cells and terminating in TEMRAs (Fig. 2g). As expected, early stages were characterized by ribosomal and translational genes (RPL10, RPL32, RPS12, TPT1), consistent with biosynthetic priming, whereas intermediate states expressed signaling and regulatory molecules including CCR7, JUNB, ZFP36, DUSP2, and IL32. Terminal effector states were marked by cytotoxic mediators such as CST7, CTSW, PRF1, and GNLY, together with effector-associated receptors (KLRG1, FCRL6) and persistence factors (LGALS1, EFHD2). The associated pathways reflected this continuum, beginning with lymphoid development and antigen processing and culminating in granzyme-mediated cytotoxicity and programmed cell death (Fig. 2g, right).

It has been well established that clonal expansion occurs in parallel with differentiation into effector and memory subsets ^10,11^. To test this, we assessed clonal architecture using V(D)J sequencing. ARIA+ patients harbored a greater burden of expanded clones than ARIA-controls, with an over-representation of medium and large clones (Fig. 2h). Linking transcriptomes to clonal size showed that larger CD8^+^ clones were increasingly enriched for effector–cytotoxic programs, including GZMB/GZMH/GNLY and the chemokine CCL4 ^16^. Regulators (EOMES, LYN) ^17,18^, NK/cytotoxic receptors (KLRC2, CMKLR1, SIGLEC17P), and antigen-presentation/adhesion genes (HLA-DQA1, ITGAM) were also elevated, and pathway analysis confirmed progressive enrichment for cell-killing functions with clonal expansion (Fig. 2g).

Integration of transcriptional data with clone size demonstrated that larger clones were enriched for effector and cytotoxic gene programs (Fig. 2i). Prominent among these were granzymes B and H (GZMB, GZMH) and granulysin (GNLY), which mediate perforin-dependent lysis of target cells ^16^. The chemokine CCL4, a CCR5 ligand that promotes recruitment of leukocytes to inflamed tissues, was also increased ^19^. Transcriptional regulators including EOMES and the Src-family kinase LYN were enriched, consistent with effector CD8 differentiation and TCR signaling modulation, ^17,18^ and expanded clones expressed cytotoxic or NK-associated receptors (KLRC2, CMKLR1, SIGLEC17P) and molecules linked to antigen presentation and adhesion (HLA-DQA1, ITGAM). Pathway analysis aligned with these observations, showing that larger clones were increasingly enriched for programs involved in cell killing and cytotoxic effector function (Fig. 2i, right).

Together these data show that ARIA+ patients exhibit remodeling of the CD8 compartment across multiple axes. Naïve cells remain unchanged in frequency and clonality, TRM-like cells maintain stable numbers but undergo selective clonal expansion, and both TEM and TEMRA subsets expand in number and clonality while acquiring progressively stronger cytotoxic programs. These findings mirror prior reports that clonally expanded CD8^+^ TEMRAs patrol the CSF in AD, enriched for cytotoxic effectors and in some cases specific for viral antigens ^20^. Similar patterns of antigen-driven clonal expansion have been described in CMV infection ^21^, multiple sclerosis ^22^, and aging-related immunosenescence ^23^, supporting the conclusion that ARIA is associated with both numerical and clonal reshaping of antigen-experienced, high-effector capacity CD8^+^ subsets. In this context, ARIA appears to reflect a coordinated metabolic, cytotoxic, and clonal reshaping of the effectorCD8^+^ pool, positioning these cells for vascular engagement and potential endothelial injury.

### Monocyte signaling and vascular ‘homing’ link CD8^+^ effectors to ARIA pathology

While these features define the intrinsic state of TEM and TEMRA cells, they raise the question of what extrinsic cues initiate and sustain such reprogramming. Because T cell differentiation and cytotoxic potential are strongly influenced by cell:cell signaling within the peripheral immune compartment, and because ARIA pathology manifests at the vascular interface, we asked if altered intercellular communication underlies these expanded TEM and TEMRA cells. To this end, GSEA analysis revealed differential (ARIA+ vs. ARIA-) enrichment of multiple terms analysis across the CD8^+^ subtypes (Fig. 3a). Pathway enrichment clustered around immune interfaces (chemotaxis, lymphocyte activation, interferon signaling, antigen presentation), vascular interfaces (BBB transport, endothelial activation, integrins, vessel morphogenesis), and neural context (APP catabolism), indicating ARIA-associated TEMRA states potentially linked to cerebrovascular injury and amyloid response.

**Figure 3.**
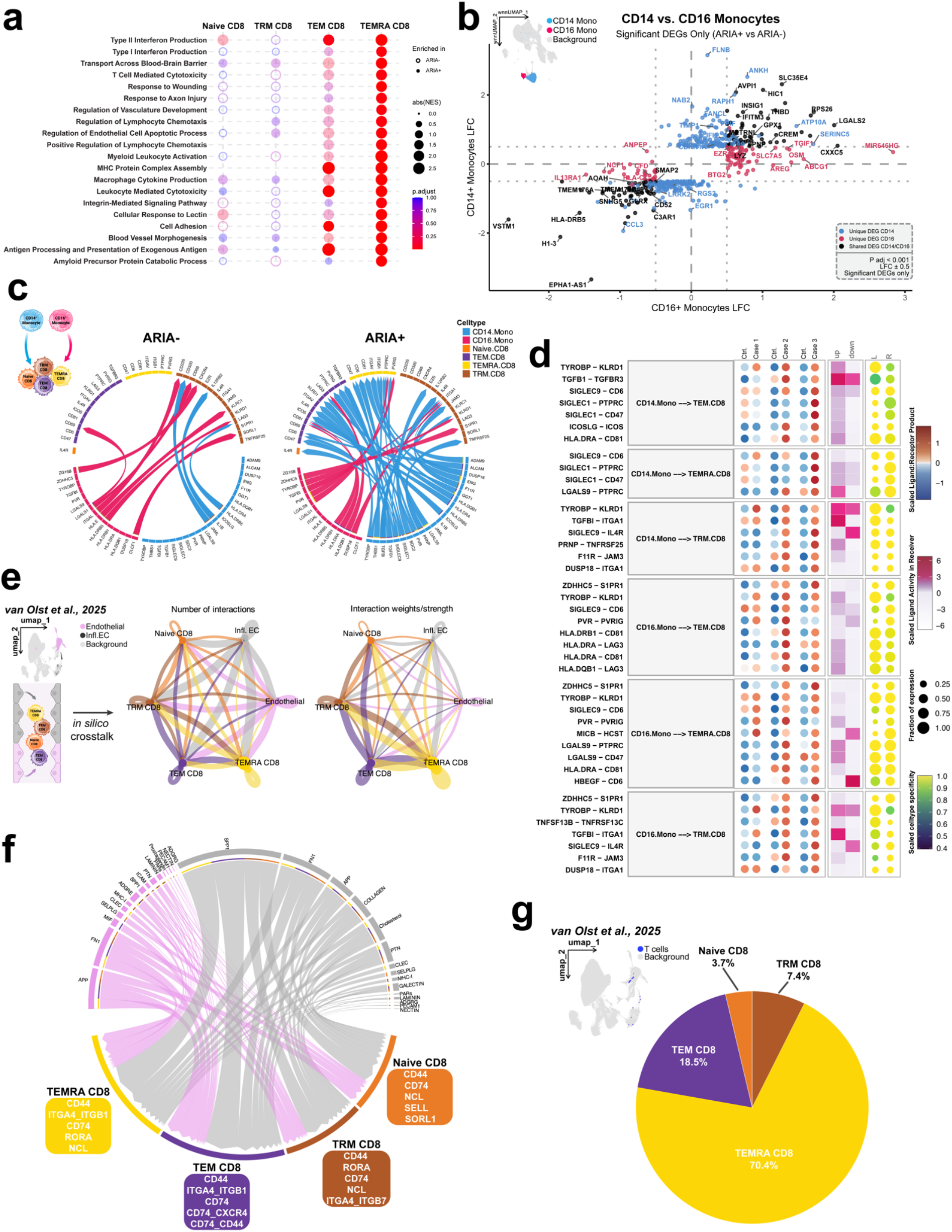
ARIA reshapes monocyte–T-cell communication and primes eKector CD8^+^ subsets for cerebrovascular engagement. **a)** Gene-set enrichment analysis (ARIA^+^ vs ARIA^−^) across CD8 subsets highlights immune-interface programs (chemotaxis, lymphocyte activation, type I/II IFN, antigen presentation), vascular-interface terms (BBB transport, endothelial activation, integrins, vessel morphogenesis), and neural-context pathways (wound response, APP catabolism). **b)** Monocyte states. CD14^+^ vs CD16^+^ monocytes show distinct transcriptional programs that are accentuated in ARIA^+^; ARIA^+^ monocytes upregulate HLA class II, adhesion molecules (ICAM1, VCAM1), and chemokines (CCL3/CCL4/CXCL10). **c)** Cell–cell communication (CellChat). TEM and TEMRA receive the strongest incoming signals from CD14^+^/CD16^+^ monocytes in ARIA^+^ samples. **d)** Recurrent ligand–receptor axes include antigen-presentation/TCR interactions, adhesion (ICAM1–LFA-1, CD58–CD2), chemokine signaling (CCL3/4–CCR5, CXCL10–CXCR3), and inhibitory circuits (PD-L1– PD-1, TIGIT–PVR). **e)** Cross-modal mapping. Blood-derived CD8 subsets projected onto a brain endothelial interaction framework (single-nucleus data from lecanemab-treated cases) show strongest connectivity for TEM/TEMRA via adhesion and chemokine receptors. **f)** Chord diagrams indicate broader and stronger inferred endothelial interactions in ARIA^+^ than ARIA^−^. **g)** Pie chart showing that TEMRA cell population accounts for most predicted cerebrovascular interactions (TEM and TRM-like intermediate; naïve minimal).

To investigate whether altered intercellular communication contributes to the expansion and effector reprogramming of CD8^+^ subsets in ARIA, we next examined ligand–receptor interactions between CD14^+^ or CD16^+^ monocytes and CD8^+^ T cells. Prior work has shown that monocytes can upregulate MHC molecules and co-stimulatory ligands, thereby shaping T-cell activation and differentiation ^24,25^. Comparing CD14^+^ and CD16^+^ monocytes revealed marked transcriptional differences that were exaggerated in ARIA+ samples (Fig. 3b). CD14^+^ monocytes showed enrichment for pathways linked to type I interferon signaling, antigen presentation, and leukocyte adhesion, while CD16^+^ monocytes preferentially expressed transcripts involved in vascular interactions, extracellular matrix remodeling, and cell killing.

Network-level analyses highlighted that TEM and TEMRA cells received most incoming signals from CD14^+^/CD16^+^ monocytes in ARIA+ samples. Chord and dot plots revealed several recurrent ligand-receptor axes, including HLA class II-CD4/TCR interactions reflecting monocyte antigen presentation to CD8^+^ cells ^26^, ICAM1-LFA1 and CD58-CD2 adhesion axes that stabilize immune synapses^27^, and chemokine signaling (CCL3/4-CCR5, CXCL10-CXCR3)^28^ pathways previously implicated in vascular homing and effector cell recruitment (Fig. 3c-d). Together these results indicate that ARIA+ monocytes not only present antigen but also create an adhesive and chemokine-rich milieu that preferentially engages TEM/TEMRAs.

Since our dataset comprises peripheral blood only, we leveraged a complementary brain single-nucleus resource to ask whether blood-derived CD8 programs mirror vascular-interacting states. Using the lecanemab (LCMB) dataset from van Olst et al., 2025^9^, we projected our CD8^+^ subsets into an endothelial-interaction framework (Fig. 3e).This analysis revealed that TEM and TEMRA cells established an *in silico* connectivity to brain endothelial clusters, through adhesion and chemokine receptor pathways similar to those identified in our monocyte analyses (ICAM1-ITGAL/ITGB2, VCAM1-ITGA4/ITGB1, CXCL10-CXCR3). Chord diagrams illustrated that ARIA+ samples showed broader and stronger inferred interactions across endothelial subtypes (Fig. 3f), reinforcing the view that these effector subsets are transcriptionally primed for vascular engagement.

Summarizing these signals, TEMRAs accounted for most predicted cerebrovascular interactions, followed by TEM and TRM-like cells, with naïve cells contributing minimally (Fig. 3g). Beyond their cytotoxic program, TEMRAs also carry the chemotaxis/adhesion repertoire that favors vascular engagement (for example, CCR5/CXCR3/CX3CR1, ITGAL/ITGB2, ITGA4/ITGB1, and CD2–CD58/ICAM1–LFA-1), which our monocyte→T-cell analyses and *in silico* endothelial mapping both prioritize. When we integrate these signatures with the cerebrovascular and Aβ-niche context reported in the van Olst study ^9^, our data support a model in which peripheral TEM/TEMRAs are transcriptionally “addressed” to the diseased vascular interface and plausibly traffic into affected tissue under inflammatory conditions.

## Discussion

This case-control study identifies a coordinated peripheral immune program associated with ARIA during lecanemab treatment. Across multiple data modalities, we observed expansion of CD8+ TEM and TEMRA subsets with metabolic reprogramming toward glycolysis, increased clonal burden, and enhanced cytotoxic programming. These intrinsic features were coupled with altered monocyte-T cell communication axes and transcriptional signatures aligned with cerebrovascular engagement. Together, these findings establish a peripheral-to-vascular immune axis that may contribute to ARIA pathophysiology and inform biomarker development for risk stratification.

The expansion of CD8^+^ TEMRA cells in ARIA+ patients extends emerging evidence linking CD8+ populations to AD pathology^29-31^. Gate et al. ^20^ identified clonally expanded CD8+ TEMRAs patrolling the CSF in AD, enriched for cytotoxic effectors and, in some cases, harboring specificity for viral antigens. Our findings parallel this observation but critically demonstrate that these changes are detectable peripherally and associate with treatment-emergent vascular pathology. The transcriptional features we observed – high expression of granzymes, perforin, and granulysin alongside inhibitory receptors and stress markers – align with established TEMRA biology in aging and chronic inflammation ^32,33^. This dual program of potent cytotoxicity and partial exhaustion suggests chronic antigenic stimulation, consistent with the “inflammaging” hypothesis where persistent low-grade inflammation drives terminal differentiation ^34^.

The unexpected expression of MHC class II molecules on ARIA+ TEMRAs warrants consideration. While CD8+ T cells classically recognize MHC class I, MHC-II expression has been documented in chronic inflammatory settings through two mechanisms: direct transcriptional upregulation in response to inflammatory signals, or acquisition from antigen-presenting cells via trogocytosis ^26,35^. Recent work has shown that HLA-DR^+^ CD8^+^ T cells correlate with disease severity with enhanced effector functions ^36,37^. In our ARIA context, this may reflect heightened activation state or increased interaction with professional antigen-presenting cells (APCs), particularly given the concurrent upregulation of antigen presentation pathways in monocytes, which is in line with prior reports of lymphocyte:monocyte cross-talk at the CNS borders^24,38^.

Importantly, similar CD8^+^ expansions characterize other CNS pathologies with vascular involvement. In multiple sclerosis, TEMRA and tissue-resident memory populations localize to perivascular and meningeal regions, where they undergo clonal expansion and contribute to lesion formation ^22,39,40^. Experimental models further demonstrate that activated CD8^+^ cells can directly injure vascular endothelium through perforin-dependent mechanisms, leading to blood-brain barrier (BBB) disruption ^41^. These precedents support a model where expanded peripheral TEMRAs in ARIA+ patients could similarly engage and damage cerebral vasculature. The metabolic shift toward glycolysis in ARIA+ CD8^+^ cells provides mechanistic insight into their functional state. Our transcription factor inference revealed downregulation of VHL, the E3 ubiquitin ligase that targets HIF-1α for degradation. Loss of VHL function stabilizes HIF-1α, driving metabolic reprogramming from oxidative phosphorylation to aerobic glycolysis, a well-characterized phenomenon in effector T cells ^42^. This metabolic state, while less efficient for ATP generation, provides rapid energy and biosynthetic precursors necessary for effector functions including cytokine production, proliferation, and cytotoxic granule synthesis ^43,44^. Recent community-based work further supports this framework by showing that peripheral CD8^+^ TEMRAs correlate with plasma markers of neuronal injury (Nf-L) and vascular:astrocytic disruption (GFAP), even in the absence of AD-specific biomarker changes ^45^. While their study did not directly link CD8^+^ TEMRAs to cognition or amyloid pathology, the alignment of TEMRA frequency with injury markers suggests that these cells act as sentinels of tissue damage. Our findings extend this concept to the vascular domain, demonstrating that CD8^+^ TEMRAs expand specifically in the setting of treatment-emergent vascular injury during ARIA.

The metabolomic validation showing elevated lactate and pyruvate with reduced TCA cycle intermediates confirms active glycolytic flux. This pattern mirrors the metabolic signature of short-lived effector cells, which prioritize immediate function over long-term survival ^46^. Scharping et al.^47^ demonstrated that sustained glycolysis in CD8+ cells correlates with enhanced cytotoxicity but reduced persistence, potentially explaining why ARIA+ TEMRAs express both effector molecules and exhaustion markers. This metabolic reprogramming likely represents both cause and consequence of effector differentiation. Cheng et al.^13^ showed that enforcing glycolysis drives CD8+ cells toward effector fates, while blocking glycolysis promotes memory formation. In the ARIA context, the combination of antigenic stimulation (likely from Aβ-antibody complexes) and inflammatory signals from activated monocytes may create a feed-forward loop where metabolic reprogramming sustains effector expansion and function.

The increased clonal burden in ARIA+ patients, particularly within TEM and TEMRA compartments, indicates antigen-driven proliferation. The correlation between clone size and cytotoxic gene expression follows established principles where TCR stimulation strength determines both proliferative capacity and effector differentiation ^10,11^. The enrichment of EOMES and TBX21 in expanded clones aligns with their roles as master regulators of CD8+ effector differentiation ^17^. The pattern of clonal expansion, minimal in naïve cells, selective in TRM-like cells, and pronounced in TEM/TEMRA, suggests progressive selection during differentiation. This mirrors observations in chronic viral infections, where CMV-specific CD8^+^ cells can comprise >10% of the total CD8^+^ pool and exhibit similar TEMRA phenotypes ^21^. In aging-related immunosenescence, such clonal expansions correlate with reduced immune diversity and increased susceptibility to novel pathogens ^23^. Whether ARIA-associated clones recognize Aβ epitopes, vascular antigens, or latent viral antigens reactivated during inflammation remains to be determined through TCR sequencing and antigen specificity testing.

The altered monocyte-T cell signaling in ARIA+ samples provides the extrinsic cues necessary for CD8^+^ activation and trafficking. The upregulation of adhesion molecules (ICAM1, VCAM1) and chemokines (CCL3/4, CXCL10) by ARIA+ monocytes creates a milieu favoring T cell recruitment and activation. These molecules mediate the classical adhesion cascade whereby circulating lymphocytes undergo selectin-mediated rolling, chemokine-triggered activation, integrin-dependent firm adhesion, and ultimately transmigration ^27,48^. The specific chemokine-receptor pairs we identified, CCL3/4-CCR5 and CXCL10-CXCR3, have established roles in neuroinflammation. CCR5^+^ T cells preferentially traffic to inflamed CNS in multiple sclerosis and HIV encephalitis ^19^, while CXCR3 engagement promotes T cell migration across the BBB in response to IFN-γ-induced CXCL10 production by astrocytes and endothelial cells ^49^. The concurrent expression of LFA-1 and VLA-4 on ARIA+ TEMRAs provides the integrin repertoire necessary for firm adhesion to activated endothelium expressing ICAM1 and VCAM1.

Our *in silico* mapping to the van Olst et al.^9^ cerebrovascular atlas provides spatial CNS context for these peripheral findings. That study demonstrated microglial activation, complement deposition, and endothelial changes at sites of Aβ clearance following immunization. Our data suggest that circulating TEMRAs are transcriptionally “pre-addressed” to engage these activated vascular niches through matching adhesion and chemokine programs. The stronger predicted interactions in ARIA+ samples indicate enhanced capacity for vascular engagement, potentially tipping the balance from successful Aβ clearance to pathological vascular injury. The model emerging from this synthesis positions ARIA as a consequence of coordinated peripheral and central immune activation. Systemic antibody administration triggers Aβ mobilization and immune complex formation, activating both circulating monocytes and CNS-resident microglia. The resulting chemokine gradients and adhesion molecule upregulation create a vascular “address” that recruits glycolytically reprogrammed, clonally expanded TEMRAs. These cells, armed with cytotoxic machinery and primed for rapid effector function, may then contribute to the endothelial injury underlying ARIA-E and ARIA-H.

These findings have potential implications for ARIA risk assessment and mitigation. The peripheral signature we identified – TEMRA expansion, metabolic shift, and clonal skewing – could form the basis for a pre-treatment screening panel. Patients with elevated baseline TEMRA frequencies or glycolytic signatures might warrant modified dosing schedules or enhanced monitoring. Serial assessment during early infusions could identify evolving risk before radiographic ARIA appears.

Therapeutically, our data may key in on several intervention points. Transient metabolic modulation to reduce glycolysis during initial antibody doses might limit TEMRA expansion without compromising long-term treatment efficacy. Alternatively, temporary blockade of specific adhesion molecules (anti-VLA4, as used in natalizumab for MS) or chemokine receptors (CCR5 antagonists) during peak risk periods could reduce CNS trafficking while preserving peripheral immunity. The timing would be critical; intervention should coincide with early Aβ mobilization but not persist long enough to impair beneficial microglial clearance mechanisms.

Critical next steps include temporal profiling across the infusion timeline to establish the kinetics of immune changes relative to ARIA onset. A larger cohort stratified by *APOE* genotype would enable robust statistical analysis and risk factor identification. Functional validation through ex vivo cytotoxicity assays, endothelial co-cultures, and chemotaxis experiments would establish causality. TCR sequencing could identify the antigens driving clonal expansion; whether Aβ epitopes, vascular proteins, or viral antigens. Finally, development of a simplified clinical assay focusing on key markers (ex. TEMRA frequency, metabolie ratios, selected chemokines) would enable prospective validation and eventual clinical implementation.

In summary, we describe here a coordinated peripheral immune program associated with ARIA during lecanemab treatment, characterized by metabolic reprogramming, clonal expansion, and altered intercellular communication in CD8^+^ effector populations. Despite limitations inherent to a small study, these findings provide biological insights into ARIA pathogenesis and establish a foundation for biomarker development. As anti-amyloid therapies become standard care, understanding and mitigating ARIA risk through precision medicine approaches will be essential for optimizing patient outcomes.

## Limitations

This study has inherent constraints that should be considered when interpreting results. The sample size of three matched pairs limits statistical power and precludes definitive conclusions about ARIA mechanisms. We cannot assess *APOE* genotype-specific effects despite ε4 carriage being the strongest known risk factor for ARIA. Importantly, the cross-sectional design captures a single timepoint, preventing determination of whether immune changes precede ARIA development or represent a consequence of vascular injury.

Additionally, our analysis relies exclusively on peripheral blood, which may not fully reflect CNS immune events. The integration with cerebrovascular data from van Olst et al. (2025) uses an external dataset from a different cohort, requiring cautious interpretation of inferred peripheral-to-brain trafficking patterns. Technical limitations include the absence of functional validation for cytotoxic capacity or adhesion properties of identified TEMRA populations. Additionally, without antigen specificity testing, we cannot determine whether clonal expansions reflect responses to Aβ, vascular antigens, or bystander activation.

The matched case-control design, while controlling for major confounders, cannot account for all variables that might influence immune profiles, including subclinical infections, medication history, or genetic factors beyond *APOE*. Finally, the heterogeneity of ARIA presentation (E versus H) in our small cohort prevents subset-specific analyses that might reveal distinct immunological mechanisms specific to each.

## Methods

### Participant identification and recruitment

Study participants were identified and consented by Norton Neuroscience Institute Memory Clinic staffmembers. Informed consent was collected under an existing IRB protocol (UofL Institutional Review Board, #42.007). Study participants were matched by sex, *APOE* genotype, ARIA status, and infusion number.

### Blood collection and processing

Blood was collected immediately prior to lecanemab infusion in BD Vacutainer EDTA-coated tubes. After collection, blood was handed off to UK staff to be transported in biohazard containers to UK labs. PBMC and plasma isolation was done using density gradient centrifugation using SepMate tubes. 5mL of plasma was removed from the top layer of the tube for use in metabolomics, lipidomics, and biomarker assays. PBMC layer was further processed with an additional wash and RBC lysis before counting and resuspension in freezing media (90% Fetal bovine Serum (VWR), 10% Dimethylsulfoxide (Invitrogen)). Cells were then allowed to cool to -80C overnight in a slow cooler (Nalgene Mr. Frosty) before transfer to LN2 for long term storage.

### Thawing and Counting of Cells

Cells were removed quickly from LN2 and immediately placed on dry ice for transfer to a 37C water bath. Cells were monitored while thawing, and when ∼90% thawed quickly decanted into pre-warmed RPMI-1640 media (Gibco) supplemented with 10% FBS and 1% Penicillin-Streptomycin cocktail. Cells were pelleted and resuspended in 1mL of DPBS supplemented with 2% FBS for counting and downstream applications. Cell counting was conducted using a hemacytometer at a dilution factor of 1:5 in Trypan Blue. Manual cell counts were verified with Denovix Cell counter.

### Sample collection and single-cell library preparation

PBMCs were isolated from six patients (Supplemental Table 1) and processed using 10x Genomics Chromium Single Cell 5’ chemistry. Simultaneous capture of transcriptomic and surface protein expression was achieved using TotalSeq-C antibody-derived tags (ADTs) targeting 137 surface proteins (Supplemental Table 2). T cell receptor sequences were captured using 10x Genomics V(D)J enrichment. Libraries were sequenced on an Illumina NovaSeq 6000 system and processed using Cell Ranger multi v9.0.1 to generate filtered feature-barcode matrices.

### Metabolomics and Lipidomics

For PBMCs, cells were pelleted as above, medium was aspirated immediately after thawing and cells were rinsed once with 500 µL ice-cold 1× DPBS and aspirated again. Eppendorf tubes containing a 1 x 10^6^ PBMC cell pellet were placed on a bed of crushed dry ice and 1mL of ice-cold 50% HPLC-grade methanol (metabolomics) or 500 µL of pre-chilled 50:50 methanol:butanol, 10 mM ammonium formate (lipidomics) was added to each tube to quench cellular metabolic activity followed by a 10 min incubation at -80°C to ensure cell lysis. After removing from the freezer, cells were collected into a microcentrifuge tube, vortexed briefly, and placed on ice until all samples were collected. The tubes were then placed on a Disruptor Genie Cell Disruptor Homogenizer (Scientific Industries) for 5 min at 3,000 rpm. Tubes were then centrifuged at 20,000 x g for 10 min at 4°C. The supernatant containing polar metabolites was transferred to a new tube and stored at -80C. On analysis day, samples were thawed on ice, centrifuged at 14,000 × g for 10 minutes at 4 °C to pellet debris, and the clear supernatant was transferred to LC-MS vials. Data were acquired on an Agilent 1290 / 6495 LC QQQ MS system.

### Quality control and data preprocessing

Individual Seurat objects were created using Seurat v5.3.0 with filtering criteria of ≥200 detected genes per cell and ≥3 cells per gene. Quality control metrics were calculated for mitochondrial, ribosomal, and hemoglobin gene content. Cells exceeding 10% mitochondrial gene expression were excluded. Putative doublets were identified and removed using scDblFinder v1.20.2, and ambient RNA contamination was corrected using decontX v1.4.1. After quality control, the dataset comprised 116,669 immune cells across 6 samples.

### Multimodal data integration and dimensionality reduction

RNA data were normalized using SCTransform with mitochondrial percentage regression, excluding ribosomal genes from variable feature selection. Data were split by sample and the top 3,000 variable genes were selected. Principal component analysis was performed retaining the first 50 components. ADT data were normalized using centered log-ratio transformation followed by scaling. All 137 ADT features were used as variable features with principal component analysis retaining the first 30 components. Cross-sample integration was performed using reciprocal principal component analysis independently for RNA and ADT modalities. The integrated representations were combined using weighted nearest neighbor analysis to generate a unified multimodal embedding. Graph-based clustering was performed on the WNN graph using the Leiden algorithm (resolution = 1.0). UMAP visualization was computed using the WNN embedding.

### Cell type annotation and classification

Major immune populations were identified using surface protein (ADT) markers with RNA expression used for additional subset refinement. T cells were identified by CD3+ and CD3E+; B cells by CD19+ and MS4A1+; classical monocytes by CD14+ (CD14+, LYZ+) and non-classical monocytes by CD16+ (FCGR3A+, CX3CR1+); NK cells by CD56+ and NCAM1+; and dendritic cells by CD123+ (pDCs), CD141+ (cDC1), or CD1c+ (cDC2) with corresponding RNA markers CLEC4C+, CLEC9A+, or CD1C+. Additional populations included naive B cells (IgD+, CD23+; IGHD+, FCER2+), memory B cells (CD27+; TNFRSF13B+), conventional NK cells (CD56+, CD16+; NCAM1+, NKG7+), CD56bright NK cells (CD56+, CD122+; IL7R+, SELL+), proliferating cells (CD71+; MKI67+), platelets (CD62P+; PPBP+, PF4+), and hematopoietic stem cells (CD34+; KIT+). Contaminating clusters were excluded prior to downstream analysis.

T lymphocytes (65,481 cells) were subset and re-integrated using identical parameters (resolution = 0.8) for fine-grained classification. CD4+ T cell subsets included naive (CD45RA+, CD62L+; CCR7+, SELL+), central memory (CD45RO+, CD127+; IL7R+, BCL2+), effector memory (CD45RO+; GZMK+), cytotoxic (GZMB+; GZMA+, PRF1+), Th2 (CD194+; GATA3+), Th17 (CD196+; CCR6+), regulatory (CD25+, TIGIT+; FOXP3+, IL2RA+), and interferon-stimulated (IFI44L+, IFIT1+, OAS1+). CD8+ T cell subsets included naive (CD45RA+, CD62L+; CCR7+, SELL+), effector memory (CD45RO+; GZMA+, CCL5+), terminally differentiated effector memory (TEMRA; CD45RA+, KLRG1+; GNLY+, GZMB+), and tissue-resident memory (CD103+; ITGAE+, CD69+). Additional T cell populations included proliferating T cells (CD71+; MKI67+), mucosal-associated invariant T cells (TCR Vα7.2+; TRAV1-2+, KLRB1+), and γ^δ^ T cells (TCR Vδ2+; TRDV2+, TRGV9+).

### TCR sequence analysis and clonal reconstruction

TCR contigs were processed using scRepertoire v2.3.4. Clonotypes were defined by identical CDR3 amino acid sequences for both α and β chains. Clonal expansion was quantified within samples and categorized by relative frequency: Rare (≤10^−4^), Small (10^−4^ to 10^−3^), Medium (10^−3^ to 10^−2^), Large (10^−2^ to 10^−1^), and Hyperexpanded (>10^−1^). TCR data were integrated with gene expression profiles, and repertoire diversity, V(D)J gene usage, and clonal overlap analyses were performed.

### Differential expression analysis

Cell type-specific differential expression analysis used Wilcoxon rank-sum tests via Seurat’s FindMarkers function. Pairwise comparisons were made between ARIA+ and ARIA-samples matched by APOE genotype and sex (minimum 10% expression threshold). Fisher’s method was employed for meta-analysis across pairwise comparisons using the test statistic χ^2^ = −2Σln(pi). Only genes detected in ≥2 independent comparisons were included. Genes were considered significantly differentially expressed with meta-analysis adjusted p-value <0.001 (FDR) and absolute mean log fold-change >0.25. Analysis was performed separately for RNA and protein expression data.

### Clonal expansion differential expression analysis

Differential expression across T cell clonal expansion states used SCP’s RunDEtest function with Wilcoxon rank-sum tests. Clonal categories (Rare, Small, Medium, Large) were compared using log fold-change threshold >1 and adjusted p-value <0.05, focusing on upregulated genes. Gene Ontology enrichment analysis was performed on clone size-specific differentially expressed genes using biological process terms, with Gene Set Enrichment Analysis to identify pathways associated with clonal expansion states.

### Pathway enrichment analysis

Gene Ontology enrichment analysis was performed using clusterProfiler v4.14.6 with org.Hs.eg.db annotation. Biological process terms were tested using over-representation analysis with Benjamini-Hochberg correction (p-value cutoff 0.05, q-value cutoff 0.2). Gene Set Enrichment Analysis was performed on pre-ranked gene lists using gseGO (minimum gene set size 10, maximum 500, p-value cutoff 0.05).

### Cytotoxic potential scoring

Cytotoxic capacity was quantified using AUCell v1.28.0 with a curated gene signature (GZMA, GZMB, GZMH, GZMK, GZMM, PRF1, GNLY, NKG7, CTSW, CST7). Area under the curve scores were computed using the top 5% of ranked genes per cell and compared across CD8+ T cell subsets.

### Cell-cell communication inference

Intercellular communication networks were reconstructed using CellChat v2.2.0 with the CellChatDB.human ligand-receptor database. Analysis was performed separately for ARIA+ and ARIA-conditions. Communication probabilities were calculated for each ligand-receptor pair between cell type pairs, with filtering requiring ≥10 cells per group. Pathway-level communication strengths were aggregated and compared between conditions.

### Pseudotime trajectory inference

CD8+ T cell differentiation trajectories were reconstructed using Slingshot v2.14.0 implemented through SCP v0.5.6. Lineages were inferred in the integrated low-dimensional space using cell type annotations as constraints. Dynamic gene expression patterns were identified along pseudotime using 200 highly variable genes.

### Gene regulatory network analysis

Cell type-specific gene regulatory networks were constructed using CellOracle v0.20.0 with the human promoter-based reference network from the CellOracle database. After random downsampling to 20,000 cells, principal component analysis was performed with automatic selection of informative components. K-nearest neighbor imputation was applied (k = 2.5% of total cells) with balanced sampling. Cell type-specific networks were constructed (α = 10) and predictive models fitted for perturbation simulation. Gene knockout simulations were performed by setting target expression to zero with 3 propagation steps. Vector field analysis used 200 nearest neighbors for transition probability estimation.

### Transcription factor activity analysis

Transcription factor activities were inferred using decoupleR v2.12.0 with the CollecTRI network obtained via get_collectri(). The Univariate Linear Model (ULM) method was applied to SCT-normalized expression data to estimate transcription factor activities based on target gene expression with minimum regulon size of 5 genes. Differential transcription factor activity analysis was performed within each cell type using t-tests comparing ARIA+ vs ARIA-samples, with a pseudocount of 2 added before log2 fold-change calculation. Results were adjusted for multiple testing using the Benjamini-Hochberg method, with significance defined as adjusted p-value <0.05 and absolute log fold-change >0.5. Transcription factors were clustered using Ward’s hierarchical clustering based on activity patterns across cell types.

### Metabolic pathway analysis

Metabolic reprogramming in CD8+ T cell subsets was assessed using selected genes from MSigDB Hallmark pathways, supplemented with additional curated genes for lactate metabolism, TCA cycle, fatty acid oxidation, and mitochondrial biogenesis. Expression data were visualized using dot plots showing mean expression, percent of cells expressing, and statistical significance determined by Wilcoxon rank-sum tests comparing ARIA+ vs ARIA-within each CD8+ subset.

### Intercellular communication network analysis

Cell-cell communication networks were analyzed using complementary approaches. For monocyte-CD8+ T cell communication within ARIA samples, MultiNicheNet v2.1.0 and CellChat v2.2.0 analyses were performed. MultiNicheNet analysis used ligand-receptor networks and regulatory matrices from the MultiNicheNet database (human v30112033). Cell type filtering required ≥10 cells per group, gene filtering used minimum sample proportion of 0.50 and fraction cutoff of 0.05. Differential expression thresholds were set to log fold-change >0.50 and p-value <0.05 without adjustment, using top 250 targets per ligand. CellChat analysis was performed independently on ARIA- and ARIA+ subsets using the CellChatDB.human database. Communication probabilities were calculated for each ligand-receptor pair between cell types, filtering interactions with <10 cells per group. Pathway-level communication strengths were computed and network centrality scores were calculated. Merged CellChat objects enabled comparative analysis between conditions with network similarity computed for functional and structural properties. To investigate potential vascular-immune interactions, NicheNet v2.2.1 analysis was performed using endothelial cells from the reference dataset as sender cells and ARIA+ CD8+ T cells as receivers. Differentially expressed genes from Fisher’s meta-analysis of TEM and TEMRA CD8+ cells served as the gene set of interest. The top 20 ligands by corrected area under precision-recall curve were selected, with ligand-target relationships inferred using weighted networks (200 targets per ligand). CellChat analysis was performed on the integrated endothelial-CD8+ dataset using identical parameters to identify communication patterns between vascular and immune cell types.

### External dataset validation

Cell type classifications were validated against published reference data ^9^ containing T cells from lecanemab-treated samples. Cross-dataset mapping was performed using k-nearest neighbor classification implemented in SCP v0.5.6. Correlation analysis used cosine similarity metrics with results displayed as annotated heatmaps. Validation focused on CD8+ T cell subset classifications to assess cross-study reproducibility. For vascular analysis, endothelial cells and inflammatory endothelial cells were subset from prefrontal cortex samples and integrated with ARIA+ CD8+ T cells. The merged object was processed via principal component analysis and SCTransform normalization to correct for sequencing depth effects.

### Data visualization and presentation

Single-cell data visualizations were generated using multiple complementary approaches. Dimensional reduction plots, expression distributions, and cell type comparisons were created using dittoSeq v1.18.0, SCP v0.5.6, and Seurat v5.3.0. Custom plots and statistical visualizations were generated using ggplot2 v3.5.2. Functional heatmaps for differential gene expression analysis were created using ComplexHeatmap v2.22.0 with expression data centered by gene across conditions (subtracting the mean expression for each gene) to highlight relative differences between ARIA+ and ARIA-samples.

### Software and packages

Data analysis was performed using R version 4.4.2. Key software packages included: Seurat v5.3.0 ^50,51^, scDblFinder v1.20.2 ^52^, decontX v1.4.1 ^9^. scCustomize (Marsh, 2021), scRepertoire v2.3.4 ^53^, clusterProfiler v4.14.6 ^54^, org.Hs.eg.db v3.20.0 (Carlson, 2024) AUCell v1.28.0 (Aibar et al., 2017), SCP v0.5.6 (Zhang, 2025), MultiNicheNet v2.1.0 ^55^, CellChat v2.2.0 ^56^, NicheNet v2.2.1 ^55^, decoupleR v2.12.0 (^57^, OmnipathR v3.15.99 ^58^, dittoSeq v1.18.0 ^59^, ggplot2 v3.5.2 (Wickham, 2016), ComplexHeatmap v2.22.0 (Gu, 2016), Slingshot v2.14.0 ^60^, NMF v0.28 ^61^

### Statistical analysis

Statistical comparisons used Wilcoxon rank-sum tests for continuous variables and chi-square tests for categorical data. Clonal expansion differences were assessed using bootstrap resampling (n = 10 iterations). Multiple testing correction used the Benjamini-Hochberg method where appropriate. All analyses were performed using the specified software packages. Statistical significance was defined as p < 0.05.

## Data Availability

All data produced in the present study are available upon reasonable request to the authors

## Supplemental data

**Supplemental Figure 1.**
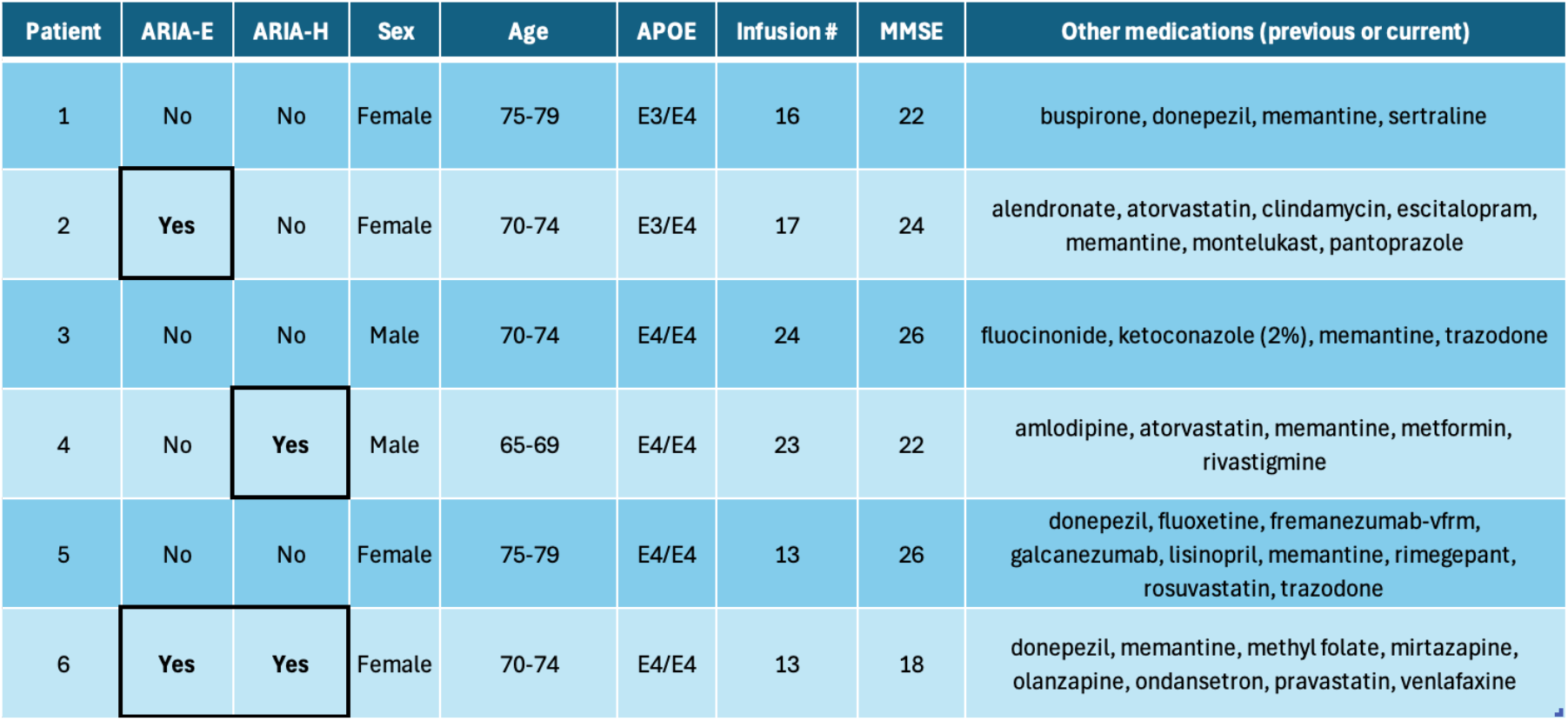
Case-control demographic information. Table showing demographic information for the three matched pairs of case-control subjects (pairs denoted by brackets). Medications includes all known treatments (past or present) during patients’ time at Norton Neuroscience Memory Clinic.

## Acknowledgements

We are deeply grateful to the patients and their caregivers who generously participated in this study. Their willingness to contribute time and effort, often under challenging circumstances, made this work possible. We thank them for their trust and commitment to advancing our understanding of Alzheimer’s disease and treatment-related complications.

